# SARS-CoV-2 infection and transmission in school settings during the second wave in Berlin, Germany: a cross-sectional study

**DOI:** 10.1101/2021.01.27.21250517

**Authors:** Stefanie Theuring, Marlene Thielecke, Welmoed van Loon, Franziska Hommes, Claudia Hülso, Annkathrin von der Haar, Jennifer Körner, Michael Schmidt, Falko Böhringer, Marcus A. Mall, Alexander Rosen, Christof von Kalle, Valerie Kirchberger, Tobias Kurth, Joachim Seybold, Frank P. Mockenhaupt, BECOSS Study Group

**Affiliations:** Institute of Tropical Medicine and International Health, Charité – Universitätsmedizin Berlin, corporate member of Freie Universität Berlin, Humboldt-Universität zu Berlin, and Berlin Institute of Health, Augustenburger Platz 1, 13353 Berlin, Germany; German Red Cross Blood Transfusion Service, Sandhofstr. 1, 60528 Frankfurt, Germany; Labor Berlin - Charité Vivantes Services GmbH, Sylter Str. 2, 13353 Berlin, Germany; Department of Pediatric Pulmonology, Immunology and Critical Care Medicine, Charité – Universitätsmedizin Berlin, corporate member of Freie Universität Berlin, Humboldt-Universität zu Berlin, and Berlin Institute of Health, Augustenburger Platz 1, 13353 Berlin, Germany; Clinical Study Center, Charité – Universitätsmedizin Berlin, corporate member of Freie Universität Berlin, Humboldt-Universität zu Berlin, and Berlin Institute of Health, Augustenburger Platz 1, 13353 Berlin, Germany; Medical Directorate, Charité – Universitätsmedizin Berlin, corporate member of Freie Universität Berlin, Humboldt-Universität zu Berlin, and Berlin Institute of Health, Augustenburger Platz 1, 13353 Berlin, Germany; Institute of Public Health, Charité – Universitätsmedizin Berlin, corporate member of Freie Universität Berlin, Humboldt-Universität zu Berlin, and Berlin Institute of Health, Augustenburger Platz 1, 13353 Berlin, Germany

**Author notes:** **Corresponding author**: Frank Mockenhaupt, **Corresponding author’s contact details**. these authors contributed equally and share first authorship (Stefanie Theuring, Marlene Thielecke, Welmoed van Loon). The members of the BECOSS study group are acknowledged at the end of the article.

## Abstract

**Background:** School attendance during the SARS-CoV-2 pandemic is intensely debated. Modelling studies suggest that school closures contribute to community transmission reduction. However, data among school-attending students and staff are scarce. In November 2020, we examined SARS-CoV-2 infections and seroreactivity in 24 randomly selected school classes and connected households in Berlin, Germany.

**Methods:** Students and school staff were examined, oro-nasopharyngeal swabs and blood samples collected, and SARS-CoV-2 infection and IgG antibodies detected by RT-PCR and ELISA. Household members performed self-swabs. Individual and institutional infection prevention and control measures were assessed. Classes with SARS-CoV-2 infection and connected household members were re-tested after one week.

**Findings:** 1119 participants were examined, including 177 primary and 175 secondary school students, 142 staff, and 625 household members. Participants reported mainly cold symptoms (19·4%). SARS-CoV-2 infection occurred in eight of 24 classes affecting each 1-2 individuals. Infection prevalence was 2·7% (95%CI; 1·2**-**5·0%; 9/338), 1·4% (0·2**-**5·1%; 2/140), and 2·3% (1·3**-**3·8%; 14/611) among students, staff and household members, respectively, including quarantined persons. Six of nine infected students were asymptomatic. Prevalence increased with inconsistent facemask use in school, way to school on foot, and case-contacts outside school. IgG antibodies were detected in 2·0% (0·8**-**4·1%; 7/347), 1·4% (0·2**-**5·0%; 2/141) and 1·4% (0·6-2·7%; 8/576), respectively. For three of nine households with infection(s) detected at cross-sectional assessment, origin in school seemed possible. After one week, no school-related, secondary infections appeared in affected classes; the attack rate in connected households was 1·1%.

**Interpretation:** These data suggest that school attendance under preventive measures is feasible, provided their rigorous implementation. In balancing threats and benefits of open *versus* closed schools during the pandemic, parents and society need to consider possible spill-overs into their households. Deeper insight is needed into the infection risks due to being a schoolchild as compared to attending school.

**Funding:** Senate of Berlin.

## Introduction

In the SARS-CoV-2 pandemic, schooling takes a central role in the public debate. The focus is on whether schools are safe to attend, whether children, adolescents, and/or schools are relevant sources of community infections, and whether school operation should be maintained, modified, or suspended.^1^ Compared to adults, SARS-CoV-2 infections in children tend to take a mild course or to stay asymptomatic, while contagiousness still is ambiguous.^2^ Remarkably, however, children and adolescents temporarily took top incidence positions in various studies in autumn 2020,^3,4^ and modelling studies suggest that closure of educational facilities significantly limits overall transmission.^5^ Nevertheless, there still is insufficient evidence as to whether schools actually drive the pandemic, or rather mirror it.^6,7^ Observational studies on the association of school closures with community transmission have yielded inconsistent results, ranging from none to substantial reduction.^8^ When considering infection risks, a distinction needs to be made between schools, students and age-typical, contextual whereabouts, e.g., public transport or after-school meetings. Limited data suggest that schools are not high-risk settings for SARS-CoV-2 transmission between students and/or staff.^2,9^ On the contrary, there is evidence that school attendance itself is not a risk factor but inconsistent mask use in school, contacts to COVID-19 cases, and gatherings outside the household.^10^ Therefore, risks need to be balanced against the detrimental impact school closures have on children and societies as to health, social equality, workforce, and economy.^11,12^

Germany experienced a strong second pandemic wave starting September 2020 and implemented a countrywide lockdown including school closures on December 16, 2020. During the preliminary peak of the second wave, we aimed at assessing SARS-CoV-2 infection and transmission in Berlin schools among schoolchildren, staff and connected household members as well as at estimating secondary infections arising from the school context.

## Methods

### Study design, setting and participants

This is a cross-sectional analysis of a longitudinal study among students and school staff from each one class in 24 schools in Berlin, and related household members. The present second round of examinations was conducted between November 2 and 16, 2020. During that time, SARS-CoV-2 transmission in Berlin (population, 3·8 million) was comparatively high: 14,514 cases were recorded, and the 7-day incidence was 185-210/100,000 inhabitants.^13^ A first round had taken place in June 2020, at low incidence.^14^ For the selection of schools, the city districts were divided into three socio-economic strata. In each stratum, two districts were randomly selected, and in these, two primary and two secondary schools. Three schools unable to participate were replaced by randomly resampled substitutes. Classes were selected amongst grades 3-5 and 9-11. We aimed at examining 20 students *per* class and ≤10 staff. In the first round, the proportion of students participating *per* class was 65% (range, 13%-96%). Hereafter, students and staff are considered index participants. With this second round, household members of index participants were also invited to participate.

The study was reviewed by the Ethics Committee of Charité–Universitätsmedizin Berlin (EA2/091/20). Informed written consent and assent was obtained from all participants and legal representatives.

### Cross-sectional data collection

Study teams visited the schools on a scheduled day. A brief medical history was obtained. Forehead temperature was scanned, and fever defined as ≥37·5°C. Oro-nasopharyngeal swabs (nerbe plus, Germany) were collected, and finger-prick blood samples taken onto filter-paper (BioSample Card, Ahlstrom Munksjö, Germany). Household members attended mobile clinics at school for symptom assessment and finger-pricking. They delivered self-collected swabs (oropharynx and nostriles), after beforehand having received instructions and swabs (CoronaOne, Germany). Participants absent due to illness or quarantine were visited at home, usually same-day. SARS-CoV-2 infection was determined by RT-PCR (GFE-Blut, Frankfurt, Germany). For anti-SARS-CoV-2-IgG, 4·75 mm dried blood spot discs were extracted in 250 µl buffer, and ELISA was performed on a EUROLabWorkstation (Euroimmun AG, Germany). In case of SARS-CoV-2 infection, health authorities were notified, participants received quarantine instructions, and during following days, they were repeatedly interviewed on health and potential infection sources. Participants completed a digital questionnaire (child, adolescent, and adult versions) two days before the study visit. Parameters assessed, spanning the preceding two weeks if appropriate, included household composition, signs and symptoms, contacts to SARS-CoV-2 positive persons, hand hygiene, physical distancing and facemask wearing.

Lastly, the school-related implementation of governmentally recommended infection prevention and control (IPC) measures was documented, including hygiene measures, distancing, absence rules at illness, ventilation, cohorting, staggering of teaching hours, and home-schooling.

### Follow-up data collection

For classes with detected SARS-CoV-2 infection, all associated students, staff and household members were re-tested after one week *via* self-sampling (CoronaOne, Germany). No re-testing was done if the positive index participant was quarantined, i.e., did not expose classmates or staff.

### Data processing and statistical analysis

Data collection was pseudonymised. On site, data was collected on paper and subsequently entered into REDCap electronic data capture tools.^15^ Descriptive analyses were segregated for primary and secondary school students, staff and household members.

We compared variables between SARS-CoV-2 infected and uninfected participants by computing proportions, odds ratios (ORs) and 95% confidence intervals (CIs). Variables of interest were socio-economic stratum, case-contacts and mask wearing in and outside school, hand washing, and transport to school/work. We used R version 3·6·3.

## Results

### Participants’ characteristics

We examined 1119 participants in 24 schools including 177 primary and 175 secondary school students, 142 staff and 625 household members. Fifty participants were visited at home because of illness or quarantine, or household members provided their swabs. Seventeen students and two staff had dropped out or withdrawn consent since June 2020. The median age of primary and secondary school students was 11 and 15 years, respectively; half were female (Table 1). Staff comprised largely mid-aged, female teachers and educators (91·2%, 114/125) in addition to facility personnel. Most household members were adults (73·8%, 461/625).

**Table 1.** Characteristics of study participants.

|  | Primary school students | Secondary school students | Staff | Household members |
| --- | --- | --- | --- | --- |
| No. | 177 | 175 | 142 | 625 |
| Age (years; median, range), n=1098 | 11·0 (9·0, 13·0) | 15·0 (14·0, 18·0) | 47·0 (28·0, 65·0) | 42·0 (2·0, 86·0) |
| Male sex (% , n/n) | 52·3% (92/176) | 45·7% (80/175) | 28·2% (40/142) | 48·8% (301/617) |
| Reported symptoms on examination day (% , n/n) |  |  |  |  |
| Any | 15·8% (28/177) | 20·1% (35/174) | 21·3% (30/141) | 19·7% (118/600) |
| Headache | 3·4% (6/177) | 8·6% (15/174) | 7·8% (11/141) | 5·0% (30/600) |
| Rhinorrhoea | 10·7% (19/177) | 8·0% (14/174) | 6·4% (9/141) | 10·3% (62/600) |
| Cough | 3·4% (6/177) | 2·9% (5/174) | 5·0% (7/141) | 6·0% (36/600) |
| Sore throat | 1·7% (3/177) | 5·2% (9/174) | 8·5% (12/141) | 5·0% (30/600) |
| Diarrhoea | 0·6% (1/177) | 1·1% (2/174) | 1·4% (2/141) | 2·0% (12/600) |
| Limb pain | 1·1% (2/177) | 0% (0/174) | 0% (0/141) | 1·3% (8/600) |
| Loss of smell or taste | 0% (0/177) | 0% (0/174) | 0% (0/141) | 1·5% (9/600) |
| Feeling feverish | 1·7% (3/177) | 0·6% (1/174) | 0·7% (1/141) | 1·2% (7/600) |
| Fever, $\geq 37.5^{\circ}\text{C}$, measured on-site | 1·7% (3/175) | 8·0% (14/174) | 2·9% (4/140) | 3·1% (18/579) |
| Any symptoms in the preceding 14 days (% , n/n) | 48·4% (45/93) | 61·0% (64/105) | 68·3% (86/126) | 55·5% (238/429) |
| Any self-reported chronic condition (% , n/n) | 6·3% (6/95) | 14·3% (15/105) | 28·0% (35/125) | 22·9% (99/433) |
| Regular medication (% , n/n) | 4·2% (4/95) | 10·7% (11/103) | 28·8% (36/125) | 25·2% (109/432) |
| SARS-CoV-2 infection (% , n/n) | 3·5% (6/171) | 1·8% (3/167) | 1·4% (2/140)* | 2·3% (14/611) |
| Anti-SARS-CoV-2 IgG positivity (% , n/n) | 1·1% (2/174) | 2·9% (5/173) | 1·4% (2/141) | 1·4% (8/576) |

\*, one staff member in quarantine, not attending school

Fever was present in 1·7%, 8·0%, and 2·9% of primary and secondary school students and staff, respectively, and any current symptom reported in 15·8%, 20·1%, and 21·3%. Leading complaints were rhinorrhoea, headache, sore throat, and cough (Table 1). Symptoms within the preceding two weeks were reported by 60·2% (195/324) of all index participants, with headache (37.3%, 121/324), sore throat (15.7%, 51/324), and rhinorrhoea (14.8%, 48/324) prevailing. Chronic conditions were stated by 10·5% of students and by 28·0% of staff, hypertension (2·6%, 13/494), lung disease (1·8%, 9/494), and obesity (1·0%, 5/494) leading.

Among household members, 3·1% were febrile at examination. Most commonly reported present symptoms (19·7%) were rhinorrhoea, cough, and sore throat (Table 1), whereas leading symptoms in the preceding two weeks (55·5%) were headache (30·5%, 131/429), tiredness (18·6%, 80/429), and rhinorrhoea (16·8%, 72/429). Most frequently stated chronic conditions (22·9%) included hypertension (4·6%, 29/624), obesity (3·7%, 23/624), and lung disease (2·1%, 13/624).

Swabs were available from 347 (98·6%) students, 142 (100%) staff, and 622 (99·5%) household members; 22 specimens were lost or did not yield a result. The electronic questionnaires had a response frequency ranging from 54·9% (614/1119) to 67·7% (758/1119) for individual items.

### School infection prevention and control measures

All schools reported the implementation of basic IPC measures such as signs on hand hygiene, soap and water in restrooms, and air ventilation at least three times a day. About half of the schools (10/22) had a hygiene commissioner. Most students (20/21 classes) and staff (22/23) reportedly adhered to hand hygiene and sneezing etiquette in more than half of the time. Three in four classes (18/24) had fixed teaching groups, but mixing with others outside was possible in almost all schools (22/24).

Students were not supposed to attend school with cold-like symptoms in 19 of 22 classes. More than half of the classes (13/22) did not provide online teaching. Two-thirds (15/24) of the schools did not have a facemask obligation *in* the classroom for students or staff, but *outside* the classroom it was obligatory for almost all (22/24).

### SARS-CoV-2 infections among students and staff

One-third (8/24) of the classes had one or two cases of SARS-CoV-2 infection detected summing up to ten cases (Table 2). These included six primary school students (two in one class, no close contact reported), three secondary students (two in one class, no close contact reported), and one secondary school staff. The resulting prevalence in school was 2·7% ([95% CI; 1·2**-**5·0%]; 9/338) among students and 0·7% among staff ([95% CI; 0·0-3·9%]; 1/140; excluding one isolated staff member who tested positive but had already tested positive a week earlier). Seven of the ten SARS-CoV-2 infected individuals were asymptomatic at testing. Of those, three developed compatible symptoms within 3-7 days, and five reported cold-like symptoms in the preceding 3-7 days. One positive staff and one positive student did not report previous, current, or later symptoms. None of the positive index participants required hospitalization.

**Table 2.** Characteristics of SARS-CoV-2 infections detected in school.

| **Index participant's school type** | **Social stratum** | **Ct-value** | **Temperature (°C)** | **Reported present symptoms** | **Symptoms before test day** | **New symptoms after test day** | **Self-reported contact to a confirmed or suspected case in preceding two weeks** | **Positive HHM (n/n tested) at cross-sectional assessment** |
| --- | --- | --- | --- | --- | --- | --- | --- | --- |
| Primary school | High | 14.1 | 37.6 | None (but febrile at examination) | 1 day before test: headache | 1 day after test: loss of smell and taste | Yes, outside school | Yes (1/1) |
| Primary school | Low | 18.9 | 36.1 | None | 6 days before test: elevated temperature, headache, fatigue for 2 days | None | None stated (other positive case in class) | No test result |
| Primary school | Low | 17.0 | 36.1 | Headache, cough | 5 days before test: headache and fever for 2 days | None | None stated (other positive case in class) | No (0/3) |
| Primary school | Low | 19.5 | 36.5 | None | 3 days before test: headache, eye pain | 4 days after test: anosmia | Yes, at school | Yes (1/3) |
| Primary school | Medium | 29.5 | 36.2 | Headache, sore throat | None | None | Yes, outside school | No (0/1) |
| Primary school | Low | 14.7 | 35.2 | None | None | None | None stated | Yes (3/5)* |
| Secondary school | High | 21.8 | 37.1 | None | None | 7 days after test: sense of taste changed | No data | Not tested ** |
| Secondary school | Medium | 27.1 | 37.4 | None | 10 days before test: cold for 7 days | None | Yes, at school (and other positive case in class) | Not tested |
| Secondary school | Medium | 23.3 | 36.6 | None | 7 days before test: sore throat | None | None stated (other positive case in class) | Not tested |
| Secondary School | Low | 24.4 | 36.1 | None | None | None | None stated | Not tested |

HHM, household member. Age and sex not reported to avoid identifiability of participants. \*, Family members tested four days after student; \*\*, One family member was not tested during cross-sectional study, but had tested positive elsewhere 8 days earlier

### Simultaneous SARS-CoV-2 infections among household members

Fourteen members of nine households tested positive in parallel to the school-based testing (prevalence, 2·3% [1·3-3·8%]; 14/611). Nine were adults, two pre-school children, and three students at non-study schools. Three family members entered the study four days delayed, were tested positive, and considered contemporaneously infected for the cross-sectional evaluation. Three of nine households were (partially) in quarantine (for three, ten, and twenty-one days), of which two households comprised a staff member (one negatively, one positively tested), and in the third one, two household members were infected. Of the nine positive households, six had no infected student or staff in school, whereas three did. For the three positive households with a positive student in school, extensive review could not solve the origin of infection.

Half (7/14) of the SARS-CoV-2 infected household members reported cold-like symptoms on the test day. Among the asymptomatic individuals, most reported symptoms previously and/or subsequently; one mid-aged adult was briefly hospitalized for oxygen substitution (Table 3).

**Table 3:** Characteristics of SARS-CoV 2 infections among household members, detected simultaneously to school survey.

| Index participant's school type; household No. | Positive index household member in school | Social stratum | Ct-value | Temperature (°C; self-measured) | Reported present symptoms | Symptoms before test day | New symptoms after test day | Self-reported contact to a confirmed or suspected case in preceding two weeks | Total no. of positive household members at cross-sectional testing (n/n tested) |
| --- | --- | --- | --- | --- | --- | --- | --- | --- | --- |
| Primary school 1) | Yes | High | 19.3 | 37.9 | None (but febrile at examination) | None | 1 day after test: fever, cough, felt very ill for 14 days | Yes | (2/2) |
| Primary school 2) | No | High | 20.4 | 36.0 | None | 5 days before test: feverish | 5 days after test: limb pain, weakness, felt very ill for 14 days | No | (1/3) |
| Primary school 3) | No | Low | 25.9 | 37.2 | Cough | None | None | None stated | (2/4) |
| Primary school 3) | No | Low | 25.2 | 36.7 | Sore throat | None | None | None stated | (2/4) |
| Primary school 4) | Yes | Low | 26.1 | 36.4 | None | 3 days before test: cold symptoms | Anosmia | None stated | (2/4) |
| Primary school 5) * | No | Low | 23.5 | 36.3 | Rhinorrhoea, cough | 3 days before test: cold symptoms for 10 days | None | Yes | (1/4) |
| Primary school 6) | Yes | Low | 11.9 | .. | None | 5 days before test: limb pain, anosmia | 2 days after test: fever, feeling very ill, after 7 days hospitalized for 5 days receiving oxygen | None stated | (4/6) |
| Primary school 6) | Yes | Low | 21.9 | .. | None | 14 days before test: cough | None | None stated | (4/6) |
| Primary school 6) | Yes | Low | 22.5 | .. | None | 14 days before test: mild cold | None | Yes | (4/6) |
| Secondary school 7) * | No* | Low | 21.6 | 36.3 | Cough | 14 days before test: fever and cough for 3 days | None | Yes | (3/3) |
| Secondary school 7) * | No* | Low | 19.4 | 36.3 | Rhinorrhoea | 10 days before test: start of cold | None | Yes | (3/3) |
| Primary school 8) * | No | Low | 21.4 | .. | Rhinorrhoea, anosmia | None | None | Yes | (2/5) |
| Primary school 8) * | No | Low | 16.9 | .. | Rhinorrhoea, cough, anosmia | Cold symptoms | None | Yes | (2/5) |
| Secondary school 9) | No | Low | 24.8 | 35.7 | None | 14 days before: mild cold | None | No data | (1/5) |

Age not reported to avoid identifiability of participants. \*, in quarantine

### SARS-CoV-2 IgG antibodies

Anti-Sars-CoV-2 IgG antibodies were present in 2·0% ([0·8-4·1%]; 7/347) of students, 1·4% ([0·2-5·0%]; 2/141) of staff, and 1·4% ([0·6-2·7%]; 8/576) of household members. Among infected participants, 9·5% (2/21) showed anti-SARS-CoV-2-IgG, and 14·3% (3/21) had borderline reactivity. Five presently uninfected index participants who had no antibodies in June 2020, did so in the present study. None was aware of previous infection. Thus, 1·1% ([0·4-2·6%]; 5/449) of students and staff passed through SARS-CoV-2 infection without noticing.

### New SARS-CoV-2 infections at follow-up after one week

For eight classes with attending SARS-CoV-2-positive index participants, students, staff, and connected household members were re-tested after one week. Students and staff of five of the eight affected classes were quarantined within a median of three days (range, 1-5) after swabbing. In three schools, only close contacts were quarantined. Of note, no school-related infection of students or staff was observed at re-testing. Yet, seven (1·8%) new infections were detected among 381 individuals associated with the affected classes and tested negative or not tested at baseline. These occurred as single events with respect to five classes, and with respect to one class, two new infections were detected. Two index participants tested positive at follow-up (Table 4). However, we specified their infections as not school-related: In the first case, a secondary school student was re-tested because of a positive staff at cross-sectional screening, but any contact was excluded. Instead, the student’s also positively tested household member had developed symptoms a few days before the student. In the second case, a staff member had been at home at the cross-sectional assessment to take care of a positively tested household member, and was tested positive at follow-up. Furthermore, five household members (four adults, one child) tested positive at follow-up. Except for the before mentioned household member of the positive index student, the remaining four had a positive child in school a week before. For two of them, we assumed SARS-CoV-2 transmission *via* a positive index participant, and for two household members, this remained unclear. In consequence, we conservatively estimated the attack rate following ten infections in eight school classes as 1·1% ([0.3-2.9]; 4/352 persons with exposed index participant at cross-sectional assessment).

**Table 4.** Characteristics of new SARS-CoV 2-positive cases at follow-up testing after 7 days.

| Index participant's school type | Index participant or household member | In index participants: case in class during cross-sectional testing? | Linked index participant positive at cross-sectional testing? | Other HHM positive at cross-sectional testing? | Infection assumed to be school-related? ** | Social stratum | Ct-value | Temperature (°C) | Present symptoms | Symptoms after testing | Cumulative no. of household members tested positive (n/n tested) + |
| --- | --- | --- | --- | --- | --- | --- | --- | --- | --- | --- | --- |
| Primary school | HHM | .. | Yes | Yes | Unclear | High | 15.8 | .. | Headache, limb pain, anosmia, cough | Cough for approx. 14 days | 3/4 |
| Primary school | HHM | .. | Yes | n.a. | Unclear | Low | 21.0 | 36.4 | None | None | 2/2 |
| Primary school | Index participant | No | .. | Yes | No | Low | 22.5 | 36.9 | Headache, feeling feverish | Headache, 1 day after test: anosmia, 3 days after: weakness | 2/4 |
| Primary school | HHM | .. | Yes | Yes | Yes | Low | 13.6 | 36.4 | Limb pain, dizziness | 2 days after test: anosmia, dizziness, weakness, cough for 1 month | 3/4 |
| Secondary School | HHM | .. | Yes | n.a. | Yes | Medium | 24.4 | 36.7 | None | None | 2/4 |
| Secondary School | Index participant | No* | .. | No | No | Low | 21.8 | 36.5 | Headache, diarrhoea | 1 day after: cold, limb pain, diarrhea, dizziness | 2/3 |
| Secondary School | HHM | .. | No | No | No | Low | 10.7 | 36.2 | Cold symptoms | Strong cold symptoms for a total of 5 days (started 2d before test) | 2/3 |

HHM, household member. Age not reported to avoid identifiability of participants. \*, No fixed class in this school subcohort; positive staff identified in school at cross-sectional testing; but no contact between those two cases, therefore not regarded as a secondary case. \*\*, Based on review. +, including those from cross-sectional testing

As to manifestation, two positive individuals were asymptomatic at retesting, whereas the others reported mainly cold-like symptoms (Table 4).

### Comparison of SARS-CoV-2 infected and non-infected participants

At cross-sectional assessment, SARS-CoV-2 infection was present in 4·7%, 1·9%, and 1·0% of classes located in the low, medium, and high socio-economic strata, respectively (high *vs*. low; OR, 4·71 [0·82-48·18]; Table 5). Nine in ten index participants stated to wear a facemask often or always at school, and their infection prevalence was 1·4%. Of those who wore masks never to sometimes, 14·3% tested positive (OR, 11·38 [2·28**-**59·64]). Similarly, 50% (8/16) of the non-affected classes and 12·5% (1/8) of the affected classes reported a facemask obligation in classroom. While contact to a suspected or confirmed COVID-19 case *in* school did not confer increased odds of infection, such contacts *outside* school tended to do so (infection prevalence, 8·3%; OR, 3·52 [0·56**-**16·27]). Lastly, infection tended to be more common in those who reported to walk to school (without other transport means; prevalence, 8·2%; OR, 3·84 [0·76**-**16·82]).

**Table 5.** Variable comparison between SARS-CoV-2 negative and positive index participants at cross-sectional survey.

|  | Negative, N=467<br>n (%) | Positive, N=11<br>n (%) | OR | 95% CI |
| --- | --- | --- | --- | --- |
| Female | 269 (97.8%) | 6 (2.2%) | 1 | -ref- |
| Male | 197 (97.5%) | 5 (2.5%) | 1.14 | (0.27-4.54) |
| Socio-economic stratum |  |  |  |  |
| - High | 193 (99.0%) | 2 (1.0%) | 1 | -ref- |
| - Medium | 151 (98.1%) | 3 (1.9%) | 1.92 | (0.22-23.18) |
| - Low | 123 (95.3%) | 6 (4.7%) | 4.71 | (0.82-48.18) |
| Contact to suspected or confirmed case at school |  |  |  |  |
| - No | 213 (96.4%) | 8 (3.6%) | 1 | -ref- |
| - Yes | 92 (97.9%) | 2 (2.1%) | 0.58 | (0.06-2.98) |
| Contact to suspected or confirmed case outside of school |  |  |  |  |
| - No | 271 (97.5%) | 7 (2.5%) | 1 | -ref- |
| - Yes | 33 (91.7%) | 3 (8.3%) | 3.52 | (0.56-16.27) |
| Mask wearing frequency at school |  |  |  |  |
| - Often to always | 273 (98.6%) | 4 (1.4%) | 1 | -ref- |
| - Never to sometimes | 30 (85.7%) | 5 (14.3%) | 11.38 | (2.28-59.64) |
| Mask wearing frequency in public |  |  |  |  |
| - Often to always | 299 (97.4%) | 8 (2.6%) | 1 | -ref- |
| - Never to sometimes | 5 (100.0%) | 0 (0.0%) | .. | .. |
| Hand washing frequency |  |  |  |  |
| - 0-1 times | 6 (85.7%) | 1 (14.3%) | 1 | -ref- |
| - 2-4 times | 92 (98.9%) | 1 (1.1%) | 0.07 | (0.00-5.97) |
| - ≥ 5 times | 206 (96.7%) | 7 (3.3%) | 0.20 | (0.02-10.70) |
| Transport to/from school/work: |  |  |  |  |
| Exclusively by foot |  |  |  |  |
| - No | 259 (97.7%) | 6 (2.3%) | 1 | -ref- |
| - Yes | 45 (91.8%) | 4 (8.2%) | 3.84 | (0.76-16.83) |
| Exclusively by bicycle |  |  |  |  |
| - No | 227 (96.6%) | 8 (3.4%) | 1 | -ref- |
| - Yes | 77 (97.5%) | 2 (2.5%) | 0.74 | (0.07-3.81) |
| Exclusively by car |  |  |  |  |
| - No | 249 (96.1%) | 10 (3.9%) | 1 | -ref- |
| - Yes | 55 (100.0%) | 0 (0.0%) | .. | .. |
| By public transport (exclusively, or in combination with other means of transport) |  |  |  |  |
| - No | 206 (97.2%) | 6 (2.8%) | 1 | -ref- |
| - Yes | 98 (96.1%) | 4 (3.9%) | 1.40 | (0.28-6.06) |

Among household members, infection was more prevalent in the low compared to the high socio-economic stratum (OR, 12·37 [2·68-114·84]), and in those who had contact to a suspected or confirmed case outside of work or school (OR, 5·76 [1·37-21·96]; data not shown).

## Discussion

Our essential results from schools during peaking SARS-CoV-2 transmission in Berlin are: In one third of the classes, one or two infections were detected, mostly asymptomatic. Connected household members in 2·3% were also infected; a school-related origin of infection was unlikely in two thirds. No secondary infections occurred in the affected classes within one week. The attack rate in households connected to positive classes was 1·1%. Infection prevalence in school was increased in case of rare facemask wearing in school, walking to school, low socioeconomic stratum, and case-contacts beyond school.

The SARS-CoV-2 prevalence of 2·7% among our student participants exceeds results of similar studies in Germany and other highly affected European countries in that period. Among >2500 students and staff in Saxony, Germany, in November 2020, 1·0% were SARS-CoV-2 infected; seroprevalence was 1·4%.^16^ A simultaneous study in Austrian students reported 0·4% SARS-CoV-2 prevalence,^17^ while in more than one-hundred English schools, 1·2% of students and 1·3% of staff were infected.^4^ During data collection, the 7-day incidence in Berlin among those aged 15-19 years exceeded that of younger ages (Figure 1). This accords with higher infection figures in secondary than in primary school students,^3^ but contrasts our findings. We cannot exclude an incidental finding; differences in hygiene and distancing might also be involved,^18^ e.g., mask wearing was not mandatory at primary schools. Only one of 140 attending school staff was infected at cross-sectional testing. This is in line with data from England, where SARS-CoV-2 infection was present in 0·4% of teachers, similar to other professions, arguing against increased infection risks among school staff.^4^ More than half of all participants reported mainly cold-like symptoms in the preceding two weeks, and about one in five on the test day. During study conduct, acute respiratory infections in Germany occurred at less than half the rate of previous years, likely due to enhanced hygiene measures.^19^ Then again, surveyed symptoms are subjective, and health consciousness might increase during a pandemic, possibly causing overestimations. Yet, seven of ten positively tested index participants were asymptomatic and would thus not have been identified by symptom-based testing. Similarly, five index participants unknowingly had developed antibodies. This stresses the potential benefit of routine testing in schools, as gradually being considered in several European countries.

**Figure 1.**
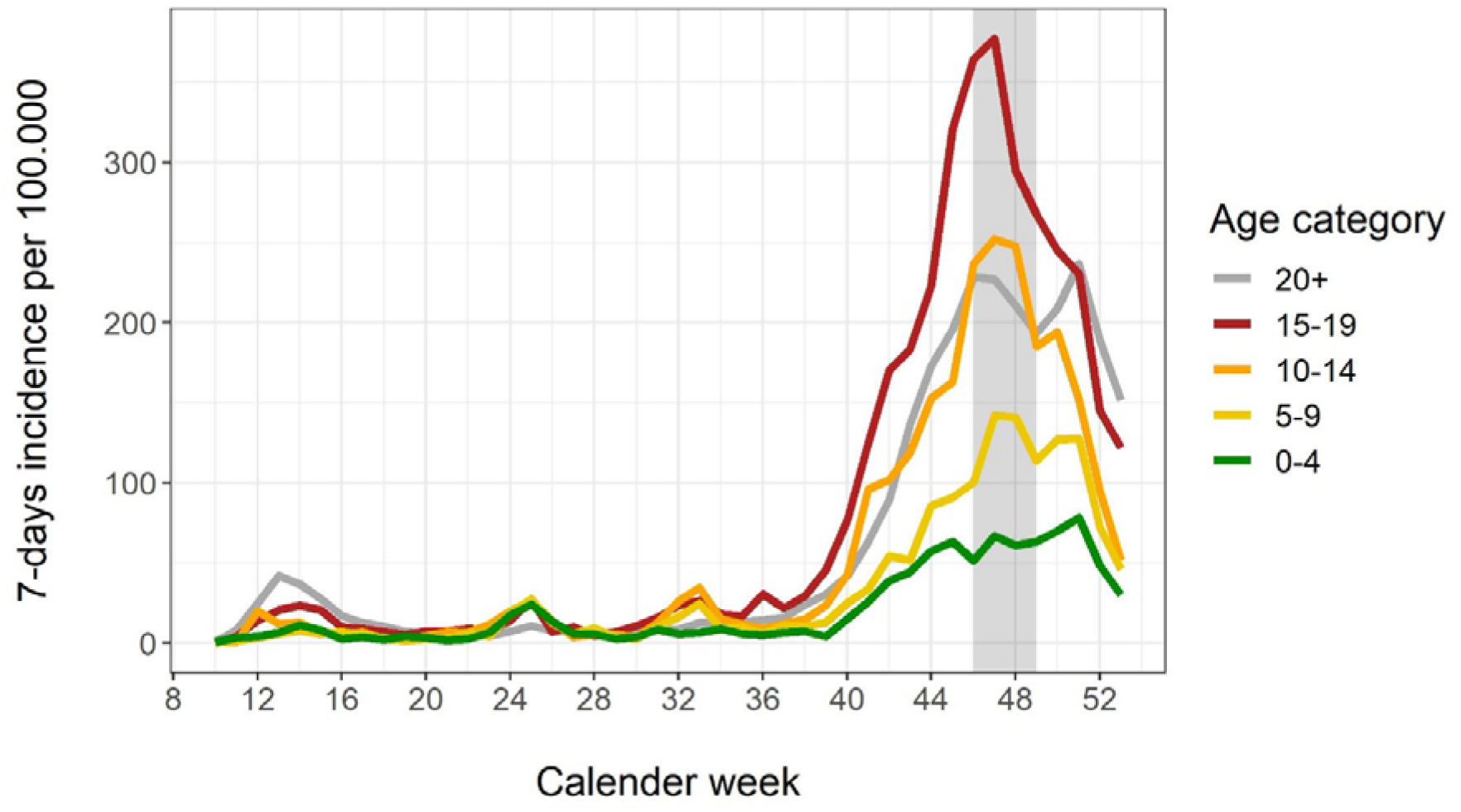
7-day incidence of recorded SARS-CoV-2 infection according to age groups in Berlin, Germany, 2020. Note. Study period indicated by grey shading. Data on PCR-confirmed SARS-CoV-2 infections, notified to local health authorities, and derived from https://daten.berlin.de/tags/covid-19-erkrankungen (German).

When comparing SARS-CoV-2 uninfected and infected index participants, the latter tended to attend school in the low socio-economic stratum. School stratum was a rough proxy disregarding intra-district variability of education, occupation and income, but also elsewhere, social disadvantage and SARS-CoV-2 infection in students were associated.^17^ Moreover, household infection clusters in our study occurred largely at low socio-economic stratum. This may reflect household crowding with insufficient distancing and isolation possibilities promoting transmission. Increased infection prevalence was observed among those who incompletely used facemasks in school. Wearing a facemask in school was not obligatory at that time, but many schools and classes nevertheless adhered to it. Prevalence was similar among participants reporting case-contacts *in* school, but for case-contacts *outside* of school, infection tended to be more prevalent. This corroborates findings from Mississippi, USA, where attending lessons was not a risk factor for SARS-CoV-2 infection among students, but inconsistent mask wearing in school, close case-contacts outside the household, and social gatherings.^10^ Lastly, prevalence was increased among those who walked to school. Lacking conclusive arguments, we suspect grouping up with friends on the way as a reason.

In the connected households, 2·3% SARS-CoV-2 prevalence was observed at cross-sectional assessment. Only for three of nine affected households, a school-link was assumed. At re-testing, no school-related secondary infection was seen among students and staff of eight affected classes, despite ongoing exposure before quarantine. For the connected households, the attack rate was 1·1%. This suggests that, even at high epidemic activity, attending lessons is not a major place of transmission if adequate IPC measures are implemented. So far, only few larger school outbreaks occurred in Germany.^20,21^ In the federal state of Rhineland-Palatinate between August and December 2020, school surveillance yielded a secondary attack rate among primary contacts of around 1%.^22^ A simultaneous investigation in neighbouring Hesse found an average secondary attack rate of 1·3% among contact persons in school.^23^ Likewise in Italy, low school prevalence and intra-school transmission prevailed up to October 2020.^24^

These findings of a rather low level of transmission in the school context are difficult to reconcile with results indicating very substantial effects of school closures. In observational US data from early 2020, school closure was associated with significant declines in COVID-19 incidence and mortality,^25^ whereas in systematic review of observational studies, effects of school closure are inconsistent.^8^ Several modelling studies - usually from the first wave of the pandemic - suggest modest to substantial associations between school closures and incidence.^5,26^ These include estimates of 40-60% reduced peak incidence,^26^ and of reducing the reproduction number by more than a third.^5^ Moreover, school closures have been associated with an overall mobility reduction of 21·6% in Switzerland.^27^ It remains difficult to disentangle the direct or indirect consequences of school closure from that of other non-pharmaceutical interventions, which were frequently implemented in parallel.^25^ For example, school closures imply less mobility, but also substantial disruptions in daily routines, particularly for parents, and altered working conditions, childcare, and social contacts. Recent evidence shows that incidence in school and population are linked.^4^ Similarly, our data suggest that most detected infections were not acquired in school. In class, students experience clear guidelines regarding preventive behaviour and respective enforcement. Such rules, e.g., facemask wearing and airing, may partially explain the rather low infection figures despite grouping in class. In contrast, during school closures, students possibly assemble in uncontrolled settings.^28^ Conceivably, shutting down educational facilities brings about transmission reductions, which are not directly attributable to attending classes and intra-school transmission, but to indirect consequences including parental behaviour. If that were true, pandemic mitigation measures would need to focus more strongly on indirect patterns, e.g., mandatory filtering masks in public, equalising public transport, and obligatory work from home wherever possible. However, there is a lack of information to delineate the respective impacts on the SARS-CoV-2 pandemic during school closures. This is all the more regrettable when considering the many harmful consequences of this measure for children and beyond.^11,12^

The strengths of our study are random selection of schools across Berlin, school-based generation of empirical data, inclusion of connected households, solid laboratory methods, and screening rather than symptom-based testing allowing for the detection of asymptomatic infections. The study is limited as to a low number of outcome events and a potential selection bias (voluntary participation). Comparative data on the prevalence of SARS-CoV-2 in the Berlin population is not available. Incomplete swabbing due to self-administration cannot completely be excluded despite illustrated instructions and PCR quality control including human *RNase P* gene co-amplification.

In conclusion, SARS-CoV-2 infection activity in Berlin schools during peak transmission appeared to be low. Secondary transmission in class was absent, and in connected households, the attack rate was around 1%. Based on our findings, we are cautiously optimistic that schooling itself does not necessarily lead to child-to-child transmission or constitute a central pandemic driver, provided that IPC measures are rigorously implemented. Continuation of our study will show whether this is true as pandemic determinants change, including vaccination coverage, herd immunity, relaxed or tightened lockdown, and viral mutations. Our findings do not exclude the possibility of school-based outbreaks, particularly at even higher transmission or enhanced viral transmissibility. Repeat screening in schools to detect also asymptomatic infections is justified by our data and should help reducing the infection burden.^29^ As a prerequisite for further, tailored measures, deeper insight is needed into the attributable fraction of infections due to being a schoolchild as compared to attending class in itself.

## Supporting information

STROBE checklist

## Data Availability

De-identified participant data and data dictionary will be available for academic institutions upon request and following approval of an analysis proposal through the Institute of Tropical Medicine and International Health, Charite - Universitaetsmedizin Berlin, for 12 months after publication of results.

## Contributors

ST, MT, FH, TK, JS, and FPM designed the study. ST, MT, WvL, CH, AvdH, JK, MAM, AR, and FPM conducted on-site examinations, interviews, and sample collection. MS and FB did laboratory examinations. ST, WvL, FH, AvdH, JK, FPM, and TK were responsible for verifying underlying data, data management and analysis. CvK, VK, and JS organized staff allocation and logistics. BECOSS study group members did data collection. ST, WvL, FH, and FPM wrote the manuscript. All authors participated in drafting the article or revising it critically for intellectual content, and approved the final version.

## Data Sharing

De-identified participant data and data dictionary will be available for academic institutions upon request and following approval of an analysis proposal through the Institute of Tropical Medicine and International Health, Charité - Universitätsmedizin Berlin, for 12 months after publication of results.

## Acknowledgements

We thank the students, school staff and families for participation, and Charité – Universitätsmedizin Berlin and the Senate of Berlin for support.

## BECOSS Study Group

Esna Bozkurt^1^, Tanja Chylla^1^, Melanie Bothmann^1^, Esra Demirtas^1^, llay Gülec^2^, Verena Haack^1^, Franziska Haniel^1^, Philipp Horn^1^, Sophia Kindzierski^1^, Mandy Kollatzsch^1^, Marco Kurzmann^1^, Sascha Lieber^1^, Elisabeth Linzbach^1^, Frederike Peters^1^, Heike Rössig^1^, Rafael Santos de Oliveira^1^, Julia Steger^1^, Zümrüt Tuncer^1^, Vanessa Voelskow^1^, Christof Wiesmann^1^

^1^ Charité - Universitätsmedizin Berlin, Berlin, Germany; ^2^ German Red Cross Blood Transfusion Service, Frankfurt, Germany

